# Limitations of PICADAR as a diagnostic predictive tool for primary ciliary dyskinesia

**DOI:** 10.1101/2025.09.22.25336363

**Authors:** Andre Schramm, Johanna Raidt, Sarah Riepenhausen, Caroline Marie Torp Nygaard, Retno Tenardi-Wenge, Tavs Qvist, Pernille Witt, Michael Storck, Heike Olbrich, Kim Gjerum Nielsen, Heymut Omran

**Affiliations:** Department of General Pediatrics, University Hospital Münster, Münster, Germany; Institute of Medical Informatics, University of Münster, Münster, Germany; Danish Primary Ciliary Dyskinesia Centre, Pediatric Pulmonary Service, Department of Pediatrics and Adolescent Medicine, Copenhagen University Hospital, Rigshospitalet, Copenhagen, Denmark; Danish PCD Centre Copenhagen, Department of Infectious Diseases, Copenhagen University Hospital, Rigshospitalet, Copenhagen, Denmark

**Keywords:** PCD, motile ciliopathy, PICADAR, predictive tool, score, test sensitivity, ciliary ultrastructure, situs inversus

## Abstract

**Background:** The Primary Ciliary Dyskinesia Rule (PICADAR) is a diagnostic predictive tool currently recommended by the European Respiratory Society (ERS) to assess the likelihood of a primary ciliary dyskinesia (PCD) diagnosis. Despite its recommendation according to the current ERS PCD diagnostic guideline, the performance of the PICADAR remains insufficiently studied.

**Methods:** We evaluated the sensitivity of PICADAR in 269 individuals with genetically confirmed PCD. Using an initial question, PICADAR rates all individuals without daily wet cough negative for PCD. PICADAR evaluates seven questions in the daily wet cough group. We here calculated test sensitivity based on the proportion of individuals scoring ≥5 points as recommended. Subgroup analyses examined the impact of laterality defects and predicted hallmark ultrastructural defects.

**Results:** 18 individuals (7%) reported no daily wet cough ruling out PCD according to PICADAR. The median PICADAR score was 7 (IQR: 5 – 9), with an overall sensitivity of 75% (202/269). Sensitivity was higher in individuals with laterality defects (95%; median score: 10; IQR 8-11) compared to those with situs solitus (61%, median score: 6; IQR 4-8; p*<0.0001). Further stratification by associated ciliary ultrastructure showed higher sensitivity in individuals with hallmark defects (83%) versus those without (59%, p*<0.0001).

**Conclusion:** The PICADAR has limited sensitivity, particularly in individuals without laterality defects or absent hallmark ultrastructural defects. Therefore, it should be used with caution as the primary factor for estimating the likelihood of PCD. Alternative predictive tools are needed, particularly for PCD individuals with normal body composition and normal ultrastructure.

## Introduction

Primary ciliary dyskinesia (PCD) is a rare inherited disorder that results from defects of the structure or function of motile cilia, which are small hair-like projections on the surface of cells. Motile cilia of airway cells move in a wave-like pattern to create a continuous flow of surrounding fluid, which is crucial for processes such as mucociliary clearance in the airways. Individuals with PCD typically experience chronic and recurrent infections of the upper and lower respiratory tract, as well as progressive decline in lung function. Many of them develop bronchiectasis over time (Shoemark et al., 2021). During early embryogenesis, motile cilia also play a functional role in determining left-right body asymmetry (Aprea et al., 2023; Despotes et al., 2024). The clinical presentation varies depending on which gene is affected and can include the presence of laterality defects and variable lung function decline (Raidt et al., 2024). So far, DNA variants in more than 50 different genes have been associated with PCD (Wallmeier et al., 2020; Despotes et al., 2024).

Given the clinical and genetic heterogeneity of PCD, diagnosing the disease is complex. The diagnostic guidelines from the European Respiratory Society (ERS) (Lucas et al., 2017) and the American Thoracic Society (ATS) (Shapiro et al., 2018) include various assessments: measurement of the nasal nitric oxide (nNO)-production rate, high-speed video microscopy (HVMA) for evaluation of the ciliary beating, immunofluorescence microscopy, transmission electron microscopy (TEM), and genetic testing. In specific cases, additional tests such as in vitro ciliogenesis may be necessary (Lucas et al., 2017). Unfortunately, no single diagnostic test can reliably diagnose PCD. This makes the diagnostic process challenging and time-consuming, as well as dependent on advanced expertise and specialized infrastructure (Nussbaumer et al., 2021). Thus, it is crucial to carefully select individuals for diagnostic testing. To aid this process, several clinical questionnaires have been developed (Martinů et al., 2021). The PrImary CiliARy DyskinesiA Rule (PICADAR) is one of the most commonly used and is recommended by the ERS guideline for PCD diagnostic (Behan et al., 2016a; Lucas et al., 2017; Castillo et al., 2023; Despotes et al., 2024). The PICADAR score comprises seven binary questions based on symptoms reported by the patients. However, it is only applicable if the prerequisite of a “daily wet cough that started in early childhood” is met. One to four points are assigned to each question. A total score of five or more suggests that further diagnostic evaluation for PCD is warranted (sensitivity 0.9) (Behan et al., 2016a). Behan et al. developed the score and proposed the cut-off in 2016 using two PCD cohorts (a derivation group (n=75), a validation group (n=80)), most of whom were diagnosed with hallmark ultrastructural defects by TEM or abnormal HVMA. However, as an increasing number of PCD-associated genes have been identified in recent years, many of which show normal ciliary ultrastructure on TEM and a normal ciliary beat pattern on HVMA, these test results have a high potential to lead to missed PCD diagnoses (Despotes et al., 2024).

Despite its widespread use, the PICADAR has not yet been evaluated in a large cohort of patients with PCD based on genetic diagnosis. Determining the test sensitivity is especially important to avoid ruling out PCD at an early stage of the diagnostic process. This study therefore aims to evaluate the sensitivity of the PICADAR in a large cohort of individuals with genetically confirmed PCD (n=269).

## Methods

### Study design

This study evaluates the sensitivity of the PICADAR in individuals with a genetically confirmed PCD diagnosis. The PICADAR score was assessed by the pulmonary teams of the University Hospital Münster and the University of Copenhagen during consultations with patients or their legal guardians. The latter provided answers on behalf of children who were too young to answer themselves.

For subgroup analyses, the study cohort was divided into different groups: PCD individuals with *situs solitus* and individuals with any type of laterality defects (e.g. *situs inversus totalis*). In a second step, subgroups were build based on genetic association with hallmark defects of the ciliary ultrastructure detectable by TEM (Raidt et al., 2024) as described by Raidt *et al*. 2024 (Raidt et al., 2024).

### Study subjects

The study included individuals from the PCD cohort of the University Hospital Münster, Germany and the University of Copenhagen, Denmark using data of the international European Reference Network (ERN) LUNG PCD registry as described before (Raidt et al., 2024). The study was approved by the Ethics Committee of the Medical Association of Westphalia-Lippe and the local ethics committee of the University of Münster (Münster, Germany; AZ 2011-270-f-S). The diagnosis of PCD was confirmed according to the ERS diagnostic guideline (Lucas et al., 2017). Only individuals with a genetically confirmed PCD diagnosis were included. Genetic variants were evaluated according to the guidelines of the American College of Medical Genetics and Genomics and the Association for Molecular Pathology (ACMG/AMP) (Richards et al., 2015), as previously described (Schramm et al., 2023; Raidt et al., 2024). Only individuals with bi-allelic (autosomal recessive inheritance), hemizygous (X-linked inheritance) or mono-allelic (autosomal dominant inheritance), disease-causing variants were included in further analyses.

### PICADAR

The PICADAR questionnaires were administered, and scores were calculated as described by Behan *et al*. 2016 (Behan et al., 2016a). If the response to the first question, “Does the patient have a daily wet cough that started in early childhood?” was negative, the PICADAR questionnaire was stopped, and a score of 0 was assigned. If the answer was positive, participants proceeded to answer the following seven questions (figure 1). If individuals were unable to answer a question, such as the second question on respiratory symptoms in the neonatal period, a ‘no’ response was assumed. In cases where the gestational age was unknown, a term birth was assumed. PICADAR scores were then calculated, and a score of five or more points was considered positive, indicating that the individua l should be further assessed for PCD (Behan et al., 2016a).

**Figure 1:**
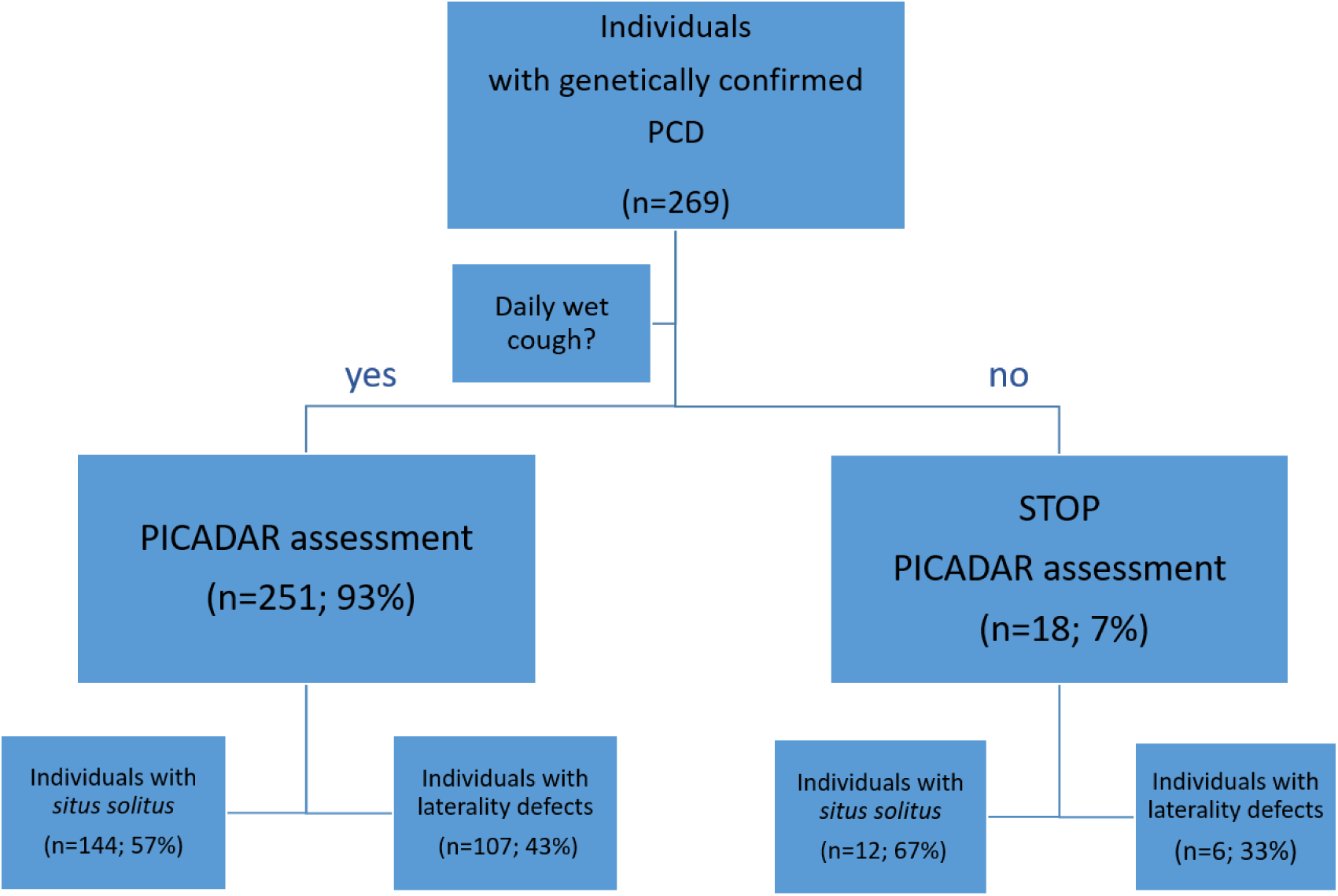
Flowchart for the determination of the PICADAR score PICADAR assessment of 269 individuals with a genetically confirmed PCD diagnosis. If the initial question regarding a daily wet cough starting in early childhood is answered with “no”, the assessment is terminated, and a score of 0 points is assigned. If the response is “yes”, the full PICADAR questionnaire is completed. Individuals are categorized into subgroups based on the presence or absence of situs abnormalities.

### Statistical Analysis

Data was extracted from the international ERN LUNG PCD registry hosted at the University of Münster (Raidt et al., 2024). Statistical analysis and visualisation were performed using R 4.5.1 with the package Tidyverse 2.0.0 (Wickham et al., 2019; R Core Team, 2025). Mann-Whitney-U tests were performed to compare the distribution of PICADAR scores and age. Fisher’s exact test was used to assess the association between positive/negative PICADAR score results and sex distribution. Boxplots were overlayed with a scatterplot of the individual scores (figure 2, 3). A horizontal line was added to illustrate the cut-off of five points. Unless otherwise stated, the median is given, with the first and third quartiles in parentheses. The significance level of alpha=0.05 has been used. In case of multiple comparisons, Bonferroni correction was applied on the calculated p-values.

**Figure 2:**
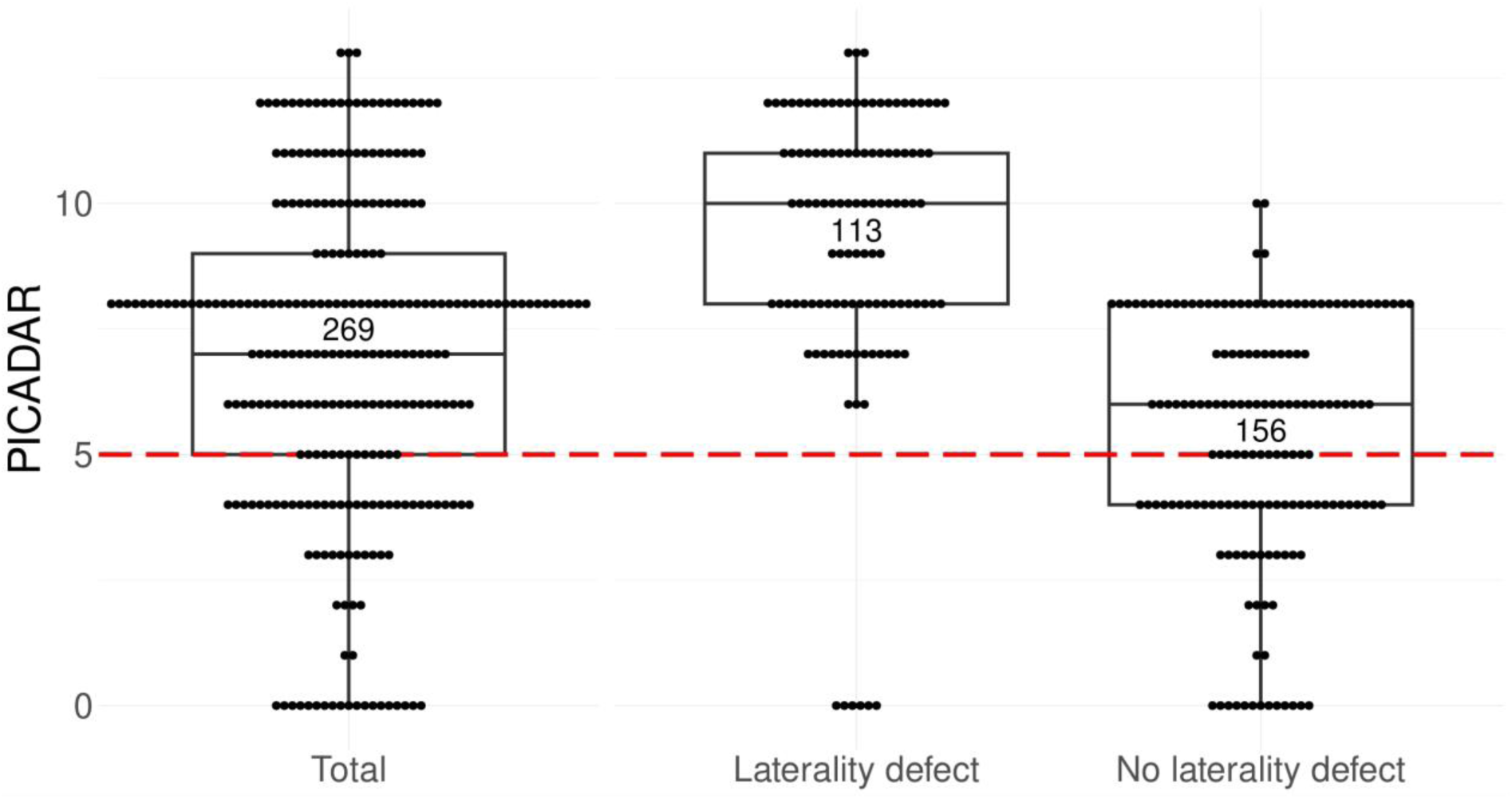
Boxplots with scatterplot of PICADAR scores among 269 individuals with PCD grouped by body composition Each point of the scatterplot represents the PICADAR score of a single individual. The dashed red line represents the threshold at 5 points to indicate the cut-off for further PCD diagnostics. In this cohort, the median PICADAR score was 7 points with 202 individuals scoring above the threshold. 18 individuals scored 0 points by negating the initial question about daily wet cough. Analysis reveals a significant difference between the groups: individuals with *situs solitus* had a median score of 6 (first quartile 4; third quartile 8), while those with laterality defects had a higher median score of 10 (first quartile 8; third quartile 11) (p*<0.0001). Within the *situs solitus* group, 39% failed the threshold (n=61/156), whereas only 5% of the group of individuals with laterality defects failed the threshold (n=6/113) all due to a negative response to the initial question (scoring 0 points).

## Results

### Study cohort

The PICADAR questionnaire was completed by 269 PCD patients. The median age of the study cohort was 26 years (first quartile 15; third quartile 40) ranging from 1 to 86 years. 134 individuals were female (50%; table 1). Disease-causing DNA variants were identified in 42 different genes associated with PCD. The most commonly affected genes in this cohort were DNAH5 (n=46, 17%) and DNAH11 (n=32, 12%). The majority of disease-causing variants follow an autosomal recessive inheritance (96%), with only a few cases of autosomal dominant inheritance (two individuals with a DNA variant in FOXJ1) or X-linked inheritance (five individuals with DNAAF6 variants, three individuals with an OFD1 variant, and one individual with a RPGR variant) (see supplementary table 1). Laterality defects such as situs inversus totalis were reported in 113 individuals (42%), whereas 156 individuals (58%) had situs solitus. There were no significant differences in age (p*=0.115) or sex distribution (p*=1) between these two groups (table 1). In addition, we grouped the whole cohort according to the genetic association with hallmark defects in ciliary ultrastructure detectable by TEM as described by Raidt et al. (Raidt et al., 2024; Shoemark et al., 2020). Disease-causing genetic variants in 185 individua ls (69%) were associated with hallmark defects in ciliary ultrastructure while variants in 85 individua ls (31%) were not associated with hallmark defects. Age and sex were similarly distributed between the two groups (p*=0.292; p*=1, respectively; table 1).

**Table 1:**
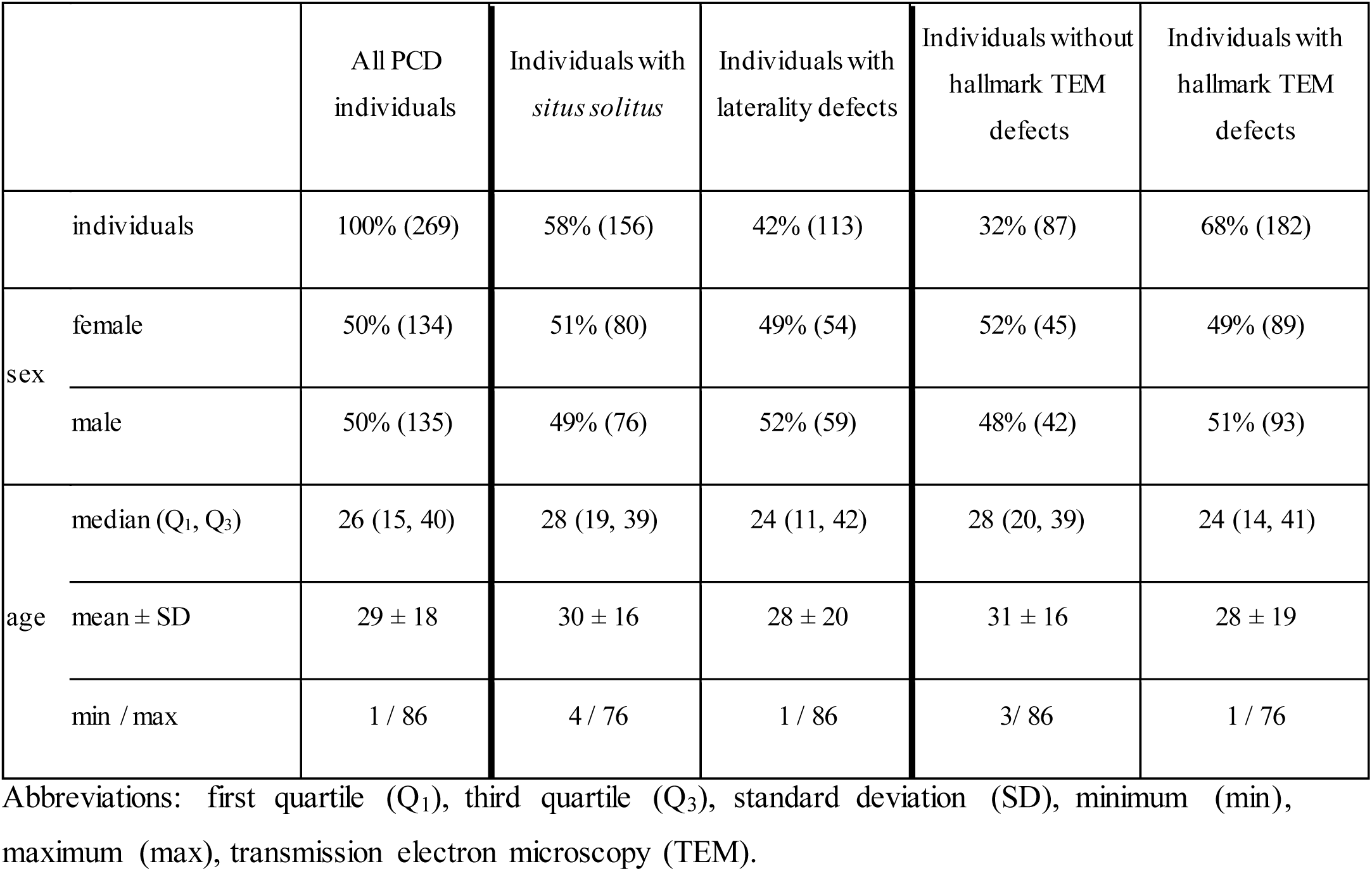
Clinical characteristics of the study cohort The table shows sex and age distribution for the total PCD cohort (n=269) and different subgroups. For subgroup analyses, the study cohort is divided into different groups: PCD individuals with situs solitus (n=156) and individuals with any type of laterality defects (e.g. situs inversus totalis; n=113) and PCD individuals with (n=87) or without hallmark defects of the ciliary ultrastructure detectable by TEM (n=182) based on genotypes as described before (Raidt et al., 2024). Unless otherwise indicated, relative numbers are given in percentages, and absolute numbers are given in parentheses.

### Analyses of the PICADAR scores

18 PCD individuals answered “no” to the question of daily wet cough (7%), resulting in a PICADAR score of 0 for all 18. Notably, six of these individuals presented with laterality defects (2%; figure 1). Individuals who reported a daily wet cough continued with the full PICADAR assessment. The distribution of responses for each question is shown in table 2. The overall median score in this cohort was 7 points (first quartile 5; third quartile 9; figure 2). Applying a threshold of 5 points to the PICADAR score results in a test sensitivity of 75% for the entire cohort (table 2). In a subgroup analysis, individuals with laterality defects had a significantly higher median PICADAR score of 10 points (first quartile 8; third quartile 11) compared to those with situs solitus, who had a median score of 6 points (first quartile 4; third quartile 8; p*<0.0001; table 2, figure 2). The subgroup of individua ls with situs solitus had a significantly lower test sensitivity of 61% (n=95/156) compared to 95% (n=107/113) in the group of individuals with laterality defects (p*<0.0001). To assess the impact of laterality defects on the performance of the PICADAR assessment, a modified PICADAR questionnaire was tested that excluded the question about laterality defects. With this modified PICADAR questionnaire, there was no significant difference between individuals with situs solitus (median 4.5; first quartile 4; third quartile 7) and individuals with laterality defects (median 6; first quartile 4; third quartile 8) (p=0.4149).

**Table 2:** Sensitivity of the PICADAR and individual components in PCD subgroups based on body composition The sensitivity of the PICADAR questionnaire and its individual components for detecting PCD is detailed in this table, presenting both relative percentages and absolute numbers in parentheses. The analysis includes the initial question as well as each component within the PICADAR questionnaire, using a score threshold of 5 points. The table shows the results for the whole study cohort (n=269), and for the subgroups of PCD individuals with *situs solitus* (n=156) and individuals with any type of laterality defects (e.g. *situs inversus totalis;* n=113)), showing a significant difference for the PICADAR score between both subgroups (p*<0.0001).

|  | **all PCD individuals**<br>**(n = 269)** | **individuals with**<br>***situs solitus***<br>**(n = 156)** | **individuals with**<br>**laterality defects**<br>**(n = 113)** |  |
| --- | --- | --- | --- | --- |
| Daily wet cough that started in early childhood? | 93% (251) | 92% (144) | 95% (107) |  |
| 1. Full term gestational age? | 87% (233) | 87% (135) | 87% (98) |  |
| 2. Neonatal chest symptoms? | 58% (157) | 60% (93) | 57% (64) |  |
| 3. Admittance to a neonatal unit? | 48% (129) | 46% (71) | 51% (58) |  |
| 4. Laterality defects? | 42% (113) | 0% (0) | 100% (113) |  |
| 5. Congenital heart defect? | 6% (17) | 5% (7) | 9% (10) |  |
| 6. Persistent perennial rhinitis? | 86% (231) | 86% (134) | 86% (97) |  |
| 7. Chronic ear or hearing symptoms? | 64% (173) | 67% (104) | 61% (69) |  |
| **PICADAR positive**<br>**(≥ 5 points)** | **75% (202)** | **61% (95)** | **95% (107)** | **p\* < 0.0001** |

In the subgroup analysis of individuals with versus without predicted hallmark defects in ciliary ultrastructure detectable by TEM, individuals with hallmark defects scored significantly higher in PICADAR (median 8; first quartile 6; third quartile 10) compared to individuals without hallmark defects (median 6; first quartile 4; third quartile 8) (p*<0.0001, figure 3). This has an impact on the sensitivity of the PICADAR between the two groups (table 3). In individuals without hallmark TEM defects, the test sensitivity of PICADAR was 59% compared to 83% in individuals with hallmark TEM defects (p*<0.0001).

**Figure 3:**
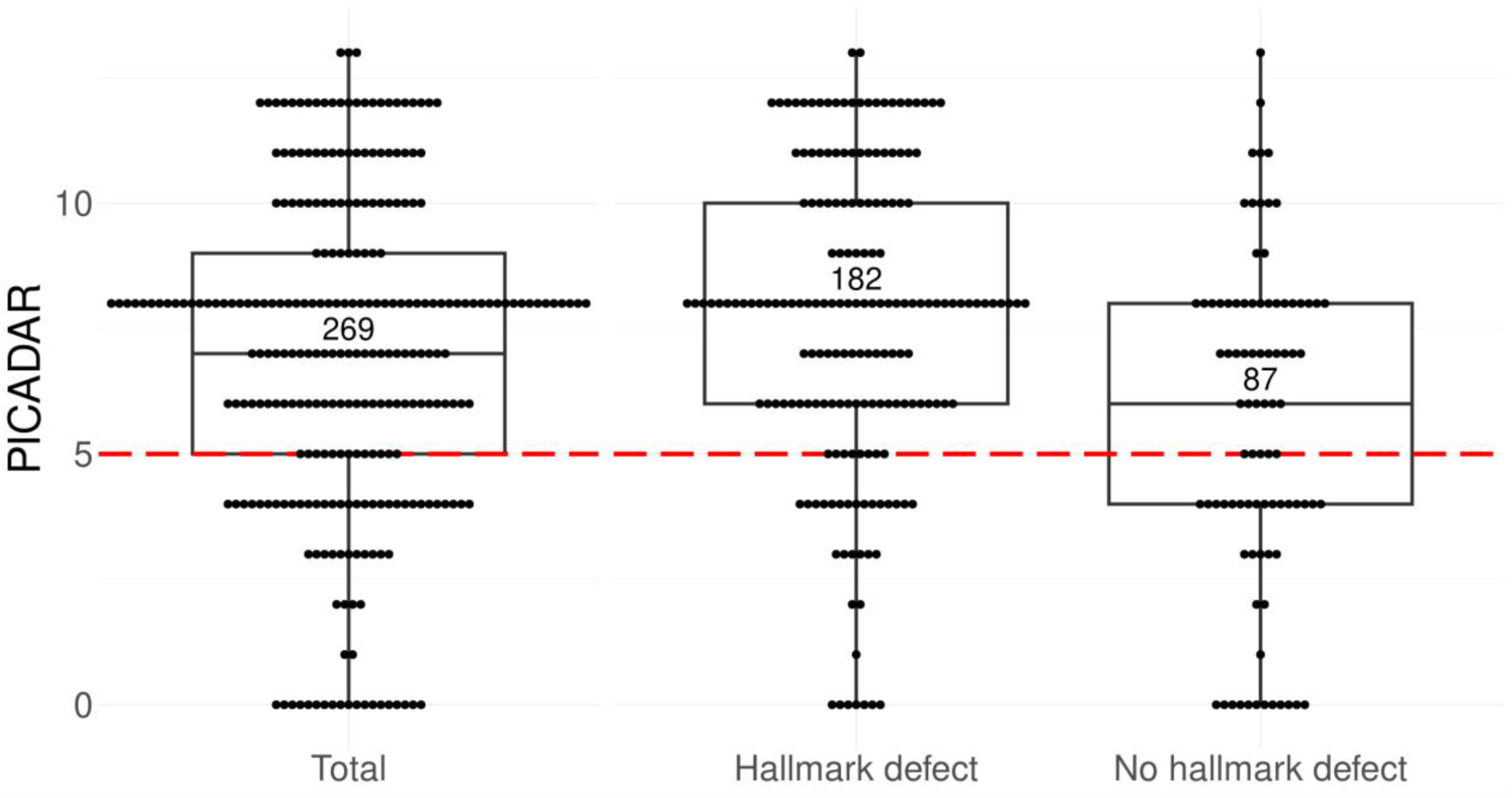
Boxplot with scatterplot of PICADAR Scores among 269 individuals with PCD grouped by hallmark TEM defects in ciliary ultrastructure based on PCD genotypes Each point of the scatterplot represents the PICADAR score of a single individual. The dashed red line represents the threshold at 5 points to indicate the cut-off for further PCD diagnostics. In this cohort, the median PICADAR score is 7 points with 202 individuals scoring above the threshold. 18 individuals scored 0 points by negating the initial question about daily wet cough. Analysis reveals a significant difference between the groups: individuals without hallmark defects in ciliary ultrastructure as detectable by TEM (Raidt et al., 2024; Shoemark et al., 2020) had a median score of 6 (first quartile 4; third quartile 8), while those with hallmark defects in ciliary ultrastructure had a higher median score of 8 (first quartile 6; third quartile 10) (p*<0.0001). Within the group without hallmark defects, 41% failed the threshold (n=36/87 individuals), whereas only 17% of the group with hallmark defects failed the threshold (n=31/182 individuals).

**Table 3:** Sensitivity of the PICADAR and individual components in PCD subgroups based on genetic association with hallmark TEM defects The sensitivity of the PICADAR questionnaire and its individual components for detecting PCD is detailed in this table, presenting both relative percentages and absolute numbers in parentheses. The analysis includes the initial question as well as each component within the PICADAR questionnaire, using a score threshold of 5 points. The table shows the results for the whole study cohort (n=269), and for the subgroups of PCD individuals with (n=87) or without hallmark defects of the ciliary ultrastructure detectable by TEM (n=182) based on genotypes as described before (Raidt et al., 2024), showing a significant difference for the PICADAR score between both subgroups (p*<0.0001).

|  | **All PCD individuals**<br>**(n = 269)** | **Individuals without**<br>**hallmark TEM defects**<br>**(n = 87)** | **Individuals with**<br>**hallmark TEM defects**<br>**(n = 182)** |  |
| --- | --- | --- | --- | --- |
| Daily wet cough that started in early childhood? | 93% (251) | 87% (76) | 96% (175) |  |
| 1. Full term gestational age? | 87% (233) | 84% (73) | 88% (160) |  |
| 2. Neonatal chest symptoms? | 58% (157) | 52% (45) | 62% (112) |  |
| 3. Admittance to a neonatal unit? | 48% (129) | 40% (35) | 52% (94) |  |
| 4. Laterality defects? | 42% (113) | 18% (16) | 53% (97) |  |
| 5. Congenital heart defect? | 6% (17) | 6% (5) | 7% (12) |  |
| 6. Persistent perennial rhinitis? | 86% (231) | 81% (70) | 89% (161) |  |
| 7. Chronic ear or hearing symptoms? | 64% (173) | 58% (50) | 68% (123) |  |
| **PICADAR positive**<br>**(≥ 5 points)** | **75% (202)** | **59% (51)** | **83% (151)** | **$p^* < 0.0001$** |

## Discussion

The diagnosis of PCD is complex, time-consuming, requiring a high level of expertise. In addition, PCD is often diagnosed late due to its non-specific symptoms (Behan et al., 2016b). Therefore, there is a demand for a predictive tool that can determine whether individuals should be referred to a specialised medical centre for PCD diagnostic assessment. A few scoring systems have been published in recent years (Martinů et al., 2021), of which PICADAR is one of the most widely used. Its use is recommended in the ERS guidelines for diagnosing PCD (Lucas et al., 2017).

Interestingly, in their original study in 2016, Behan et al. found a test sensitivity for the PICADAR of 90% (derivation group, PCD n=75) to 86% (validation group, PCD n=80) using a cut-off value of 5 points. The study cohort of PCD individuals was mainly diagnosed by TEM and HVMA. As both methods have inherent limitations and can detect only certain forms of PCD, it is very likely that individuals with other PCD variants were excluded from the PCD cohort. In addition, it is likely that their disease control group contained PCD individuals, because they only used TEM and HVMA to exclude PCD. However, we have recently shown that there are PCD types caused by defects of the C1d projection that have normal ultrastructure and normal HVMA (Wohlgemuth et al., 2024). Given the discovery of an increasing number of PCD genes that are associated with normal ciliary ultrastructure since PICADAR was introduced (Raidt et al., 2024), it is crucial to evaluate the PICADAR in a genetically defined PCD cohort. In our cohort of 269 individuals with a genetically confirmed PCD, the PICADAR score has a sensitivity of only 75%, when an identical cut-off is used. The subgroup analysis in our study showed that the PICADAR performed worse in individuals with PCD and a normal body composition (61%). Using the PICADAR as a clinical decision support tool, many of these individuals would not have been referred for PCD diagnostic work-up. However, 95% of individuals with laterality defects would be eligible for further PCD testing. Our data show that these differences in test sensitivity mainly result from the high weighting given to the fourth item, which relates to laterality defects. When this item is neglected, PICADAR performs similarly in both subgroups. Interestingly, the prevalence of laterality defects in the derivation group (44%, n=33/75) and validation group (44%, n=41/93) in the work from Behan et al. was similar to the prevalence in our cohort and therefore does not explain the observed difference in PICADAR sensitivity (Raidt et al., 2024). A genotype-phenotype correlation may be a relevant confounding factor of the PICADAR. The diagnostic algorithm implemented by Behan et al. primarily identifies PCD variants with hallmark ciliary ultrastructural defects, identifiable by TEM. These PCD variants are typically associated with situs abnormalities. Consequently, PCD variants lacking such hallmark defects were underrepresented or absent from their cohort, potentially leading to an overestimation of the sensitivity of the PICADAR reported by the colleagues. Including such cases could have resulted in substantially different sensitivity estimates.

While Behan et al. did not report these numbers in their UK derivation and validation groups, a recent European PCD registry study analysed data from 1,236 genotyped PCD individuals and found a high prevalence of hallmark ultrastructural ciliary defects in the UK cohort (n=153, 79% hallmark ultrastructural defects) (Raidt et al., 2024). In contrast, countries such as Germany and Denmark, where our cohort is based, showed lower prevalences (Germany: n=321, 69% hallmark ultrastructural defects; Denmark: n=91, 68% hallmark ultrastructural defects). Additionally, the prevalence of laterality defects varied between countries. In the UK cohort (n=153), laterality defects were observed in 45% of individuals, while 41% had a normal body composition (14% unknown situs status). In Germany (n=321), the prevalence of laterality defects was lower (37%), with 61% showing normal body composition (2% unknown). In Denmark, the prevalence of laterality defects was similar compared to Germany (41%), with 58% showing normal body composition (1% unknown). Other countries, such as Turkey, had an even lower prevalence (n=47; 28% laterality defects, 72% normal body composition). Interestingly, Raidt et al. reported a much higher prevalence of laterality defects in the large cohort of PCD individuals with hallmark ultrastructural defects detectable by TEM (n=894; 51% laterality defects, 45% normal body composition, 3% unknown) than in the group without hallmark defects (n=342; 18% laterality defects, 79% normal body composition and 4% unknown) (Raidt et al., 2024).

Thus, we considered that PICADAR may be less efficient in populations characterised by a low prevalence of genetic variants that cause laterality defects and/or leading to hallmark ciliary ultrastructure defects. This may be due either to regional factors (such as in Turkey) or to methodological limitations (Behan et al., 2016a). In our study, we present the results of 87 PCD individuals with genetic defects associated without ultrastructural hallmark defects detectable by TEM (32%; see supplementary table 1). Our data confirm that the PICADAR questionnaire performs poorly in this subgroup. If PICADAR is applied strictly, only 59% of PCD individuals whose disease-causing genetic variants are not associated with hallmark TEM defects would be referred for further PCD diagnostics. In contrast, 83% of individuals with hallmark TEM defects would receive a positive result in the PICADAR assessment.

Interestingly, only a few other studies assessed the sensitivity of the PICADAR questionnaire. Martinu et al. evaluated 67 PCD individuals and reported a sensitivity of 87%, although using a higher threshold of 6 points (Martinů et al., 2021). The PCD diagnosis was primarily based on abnormal HVMA findings, which results in a high number of individuals with abnormal ultrastructure (n=50, 75%) probably similar to the original PICADAR cohort. To date, only two rather small studies with 25 (Palmas et al., 2020) and 36 individuals (Chiyonobu et al., 2022) evaluated the PICADAR score in PCD cohorts where genetic testing was done independent of HVMA or TEM findings. These smaller studies show a sensitivity of 76% and 78% for the PICADAR similar to our findings. Interestingly, Chiyonobu et al. also showed that individuals with situs solitus had a lower median PICADAR of 5.8 compared to 10.2 in individuals with situs inversus (Chiyonobu et al., 2022). Although test sensitivity was not calculated in the original article, our re-analysis of the Japanese data showed a similar low sensitivity comparable to our findings, with 67% sensitivity for the situs solitus group compared to 100% for the situs inversus group.

Based on our findings, we recommend that the PICADAR score should be used cautiously when prioritizing individuals for PCD testing. We suggest that PCD testing should be performed for all individuals with laterality defects and chronic airway disease, regardless of whether they report a wet cough since early childhood. This procedure prevents the exclusion of individuals with PCD and laterality defects based on the results of the PICADAR (in our cohort, six individuals were excluded). In addition, we highlight a research need. For the remaining individuals with PCD and normal situs composition, clinicians need a novel tool to estimate the likelihood of PCD. Unfortunately, our findings demonstrate that PICADAR does not meet the required level of accuracy (only 61% detection rate) for this subgroup. A novel predictive tool should be developed and evaluated in a large, genetically well-defined PCD cohort. This might be best achieved in a multicentre, multinational study, e.g. using the International ERN LUNG PCD registry (Raidt et al., 2024). The diagnosis of PCD is complex, time - consuming, requiring a high level of expertise. In addition, PCD is often diagnosed late due to its non-specific symptoms (Behan et al., 2016b). Therefore, there is a demand for a predictive tool that can determine whether individuals should be referred to a specialised medical centre for PCD diagnostic assessment. A few scoring systems have been published in recent years (Martinů et al., 2021), of which PICADAR is one of the most widely used. Its use is recommended in the ERS guidelines for diagnosing PCD (Lucas et al., 2017).

Interestingly, in their original study in 2016, Behan *et al*. found a test sensitivity for the PICADAR of 90% (derivation group, PCD n=75) to 86% (validation group, PCD n=80) using a cut-off value of 5 points. The study cohort of PCD individuals was mainly diagnosed by TEM and HVMA. As both methods have inherent limitations and can detect only certain forms of PCD, it is very likely that individuals with other PCD variants were excluded from the PCD cohort. In addition, it is likely that their disease control group contained PCD individuals, because they only used TEM and HVMA to exclude PCD. However, we have recently shown that there are PCD types caused by defects of the C1d projection that have normal ultrastructure and normal HVMA (Wohlgemuth et al., 2024). Given the discovery of an increasing number of PCD genes that are associated with normal ciliary ultrastructure since PICADAR was introduced (Raidt et al., 2024), it is crucial to evaluate the PICADAR in a genetically defined PCD cohort. In our cohort of 269 individuals with a genetically confirmed PCD, the PICADAR score has a sensitivity of only 75%, when an identical cut-off is used. The subgroup analysis in our study showed that the PICADAR performed worse in individuals with PCD and a normal body composition (61%). Using the PICADAR as a clinical decision support tool, many of these individuals would not have been referred for PCD diagnostic work-up. However, 95% of individuals with laterality defects would be eligible for further PCD testing. Our data show that these differences in test sensitivity mainly result from the high weighting given to the fourth item, which relates to laterality defects. When this item is neglected, PICADAR performs similarly in both subgroups. Interestingly, the prevalence of laterality defects in the derivation group (44%, n=33/75) and validation group (44%, n=41/93) in the work from Behan *et al*. was similar to the prevalence in our cohort and therefore does not explain the observed difference in PICADAR sensitivity (Raidt et al., 2024). A genotype-phenotype correlation may be a relevant confounding factor of the PICADAR. The diagnostic algorithm implemented by Behan *et al*. primarily identifies PCD variants with hallmark ciliary ultrastructural defects, identifiable by TEM. These PCD variants are typically associated with situs abnormalities. Consequently, PCD variants lacking such hallmark defects were underrepresented or absent from their cohort, potentially leading to an overestimation of the sensitivity of the PICADAR reported by the colleagues. Including such cases could have resulted in substantially different sensitivity estimates.

While Behan *et al*. did not report these numbers in their UK derivation and validation groups, a recent European PCD registry study analysed data from 1,236 genotyped PCD individuals and found a high prevalence of hallmark ultrastructural ciliary defects in the UK cohort (n=153, 79% hallmark ultrastructural defects) (Raidt et al., 2024). In contrast, countries such as Germany and Denmark, where our cohort is based, showed lower prevalences (Germany: n=321, 69% hallmark ultrastructural defects; Denmark: n=91, 68% hallmark ultrastructural defects). Additionally, the prevalence of laterality defects varied between countries. In the UK cohort (n=153), laterality defects were observed in 45% of individuals, while 41% had a normal body composition (14% unknown situs status). In Germany (n=321), the prevalence of laterality defects was lower (37%), with 61% showing normal body composition (2% unknown). In Denmark, the prevalence of laterality defects was similar compared to Germany (41%), with 58% showing normal body composition (1% unknown). Other countries, such as Turkey, had an even lower prevalence (n=47; 28% laterality defects, 72% normal body composition). Interestingly, Raidt *et al*. reported a much higher prevalence of laterality defects in the large cohort of PCD individuals with hallmark ultrastructural defects detectable by TEM (n=894; 51% laterality defects, 45% normal body composition, 3% unknown) than in the group without hallmark defects (n=342; 18% laterality defects, 79% normal body composition and 4% unknown) (Raidt et al., 2024).

Thus, we considered that PICADAR may be less efficient in populations characterised by a low prevalence of genetic variants that cause laterality defects and/or leading to hallmark ciliary ultrastructure defects. This may be due either to regional factors (such as in Turkey) or to methodological limitations (Behan et al., 2016a). In our study, we present the results of 87 PCD individuals with genetic defects associated without ultrastructural hallmark defects detectable by TEM (32%; see supplementary table 1). Our data confirm that the PICADAR questionnaire performs poorly in this subgroup. If PICADAR is applied strictly, only 59% of PCD individuals whose disease-causing genetic variants are not associated with hallmark TEM defects would be referred for further PCD diagnostics. In contrast, 83% of individuals with hallmark TEM defects would receive a positive result in the PICADAR assessment.

Interestingly, only a few other studies assessed the sensitivity of the PICADAR questionnaire. Martinu *et al*. evaluated 67 PCD individuals and reported a sensitivity of 87%, although using a higher threshold of 6 points (Martinů et al., 2021). The PCD diagnosis was primarily based on abnormal HVMA findings, which results in a high number of individuals with abnormal ultrastructure (n=50, 75%) probably similar to the original PICADAR cohort. To date, only two rather small studies with 25 (Palmas et al., 2020) and 36 individuals (Chiyonobu et al., 2022) evaluated the PICADAR score in PCD cohorts where genetic testing was done independent of HVMA or TEM findings. These smaller studies show a sensitivity of 76% and 78% for the PICADAR similar to our findings. Interestingly, Chiyonobu *et al*. also showed that individuals with *situs solitus* had a lower median PICADAR of 5.8 compared to 10.2 in individuals with *situs inversus* (Chiyonobu et al., 2022). Although test sensitivity was not calculated in the original article, our re-analysis of the Japanese data showed a similar low sensitivity comparable to our findings, with 67% sensitivity for the *situs solitus* group compared to 100% for the *situs inversus* group.

Based on our findings, we recommend that the PICADAR score should be used cautiously when prioritizing individuals for PCD testing. We suggest that PCD testing should be performed for all individuals with laterality defects and chronic airway disease, regardless of whether they report a wet cough since early childhood. This procedure prevents the exclusion of individuals with PCD and laterality defects based on the results of the PICADAR (in our cohort, six individuals were excluded). In addition, we highlight a research need. For the remaining individuals with PCD and normal situs composition, clinicians need a novel tool to estimate the likelihood of PCD. Unfortunately, our findings demonstrate that PICADAR does not meet the required level of accuracy (only 61% detection rate) for this subgroup. A novel predictive tool should be developed and evaluated in a large, genetically well-defined PCD cohort. This might be best achieved in a multicentre, multinational study, e.g. using the International ERN LUNG PCD registry (Raidt et al., 2024).

## Ethics statement

This research project was approved by the Ethics Committee of the Medical Association of Westphalia - Lippe and the University of Muenster (reference number: 2015-104-f-S). Signed and informed consent was obtained from participating individuals, legal guardians and/or relatives by protocols approved by the Institutional Ethics Review Board of the University Muenster and collaborating institutions.

## Data Availability

All data produced in the present study are available upon reasonable request to the authors

## Acknowledgments

We thank the PCD-affected individuals and their families for their participation. We thank S. Helms and M. Tekaat (Department of General Pediatrics, University Children’s Hospital Muenster, Muenster, Germany) for excellent technical and organisational assistance and P. Pennekamp, NT. Loges as well as B. Dworniczak for excellent support in diagnosis of PCD individuals. We thank the Medical Data Integration Center (MeDIC) at the Institute of Medical Informatics at the University Münster for their support with research data management (funded by the Federal Ministry of Research, Technology and Space (BMFTR; grant number 01KX2121).

## Supplementary Material

**Supplementary table 1:** Affected genes in mutant PCD individuals *Genes that are associated with pathognomonic ciliary ultrastructure defects detectable by transmission electron microscopy (hallmark defects) according to Shoemark *et al*. (Shoemark et al., 2020).

| Affected genes | Number of PCD individuals |
| --- | --- |
| *CCDC103* * | 6 |
| *CCDC39* * | 7 |
| *CCDC40* * | 27 |
| *CCNO* | 1 |
| *CFAP221* | 1 |
| *CFAP298* * | 2 |
| *CFAP300* * | 3 |
| *CFAP45* | 1 |
| *CFAP46* | 1 |
| *CFAP54* | 1 |
| *CFAP74* | 1 |
| *DNAAF1* * | 3 |
| *DNAAF2* * | 3 |
| *DNAAF3* * | 6 |
| *DNAAF4* * | 3 |
| *DNAAF5* * | 1 |
| *DNAAF6* * | 5 |
| *DNAAF11* * | 3 |
| *DNAH11* | 32 |
| *DNAH5\** | 46 |
| *DNAH9\** | 2 |
| *DNAI1\** | 26 |
| *DNAI2\** | 4 |
| *DRC1* | 2 |
| *DRC2* | 3 |
| *FOXJ1* | 2 |
| *GAS8* | 1 |
| *HYDIN* | 13 |
| *IFT74* | 2 |
| *NEK10* | 4 |
| *ODAD1\** | 7 |
| *ODAD2\** | 6 |
| *ODAD3\** | 3 |
| *ODAD4\** | 4 |
| *OFD1* | 3 |
| *RPGR* | 1 |
| *RSPH1* | 5 |
| *RSPH4A* | 5 |
| *RSPH9* | 5 |
| *SPAG1\** | 4 |
| *SPEF2* | 3 |
| *ZMYND10\** | 11 |

